# Impact of the COVID-19 Pandemic on the Short-term Course of Obsessive-Compulsive Disorder

**DOI:** 10.1101/2020.07.26.20162495

**Authors:** Lavanya P Sharma, Srinivas Balachander, Abel Thamby, Mahashweta Bhattacharya, Chethana Kishore, Vandita Shanbhag, TS Jaisoorya, Janardhanan C Narayanaswamy, Shyam Sundar Arumugham, YC Janardhan Reddy

## Abstract

**Background:** There is an understandable concern that obsessive-compulsive disorder (OCD) may worsen during the COVID-19 pandemic, but there is little empirical data. We report the impact of COVID-19 pandemic on the short-term course of OCD. We also assessed for predictors of relapse and emergence of COVID–19–themed obsessive–compulsive symptoms.

**Methods:** A cohort of patients with a primary diagnosis of OCD (n=240) who were on regular follow-up at a tertiary care specialty OCD Clinic in India were assessed telephonically, about 2 months after the declaration of the pandemic (‘pandemic’ cohort). Data from the medical records of an independent set of patients with OCD (n=207) who were followed–up during the same period, one year prior, was used for comparison (historical controls).

**Results:** The ‘pandemic’ group and historical controls did not differ in the trajectories of the Yale-Brown Obsessive-Compulsive Scale (YBOCS) scores (Chi-square for likelihood-ratio test of the Group × Time interaction = 2.73, p= 0.255) and relapse rate [21% vs 20%, adjusted odds ratio = 0.81 (95% CI 0.41 -1.59, p=0.535]. Pre-existing contamination symptoms and COVID-19-related health anxiety measured by the COVID-Threat Scale did not predict relapse. Only a small proportion of patients (6%) reported COVID-19-themed obsessive-compulsive symptoms.

**Limitations:** Follow-up 2 months after pandemic declaration may be too early understand the true impact.

**Conclusions:** The COVID-19 pandemic, at least in the short-run, did not influence the course of illness in those who were on medications. It would be pertinent to evaluate the long-term impact of the pandemic on the course of OCD.

**Highlights:** 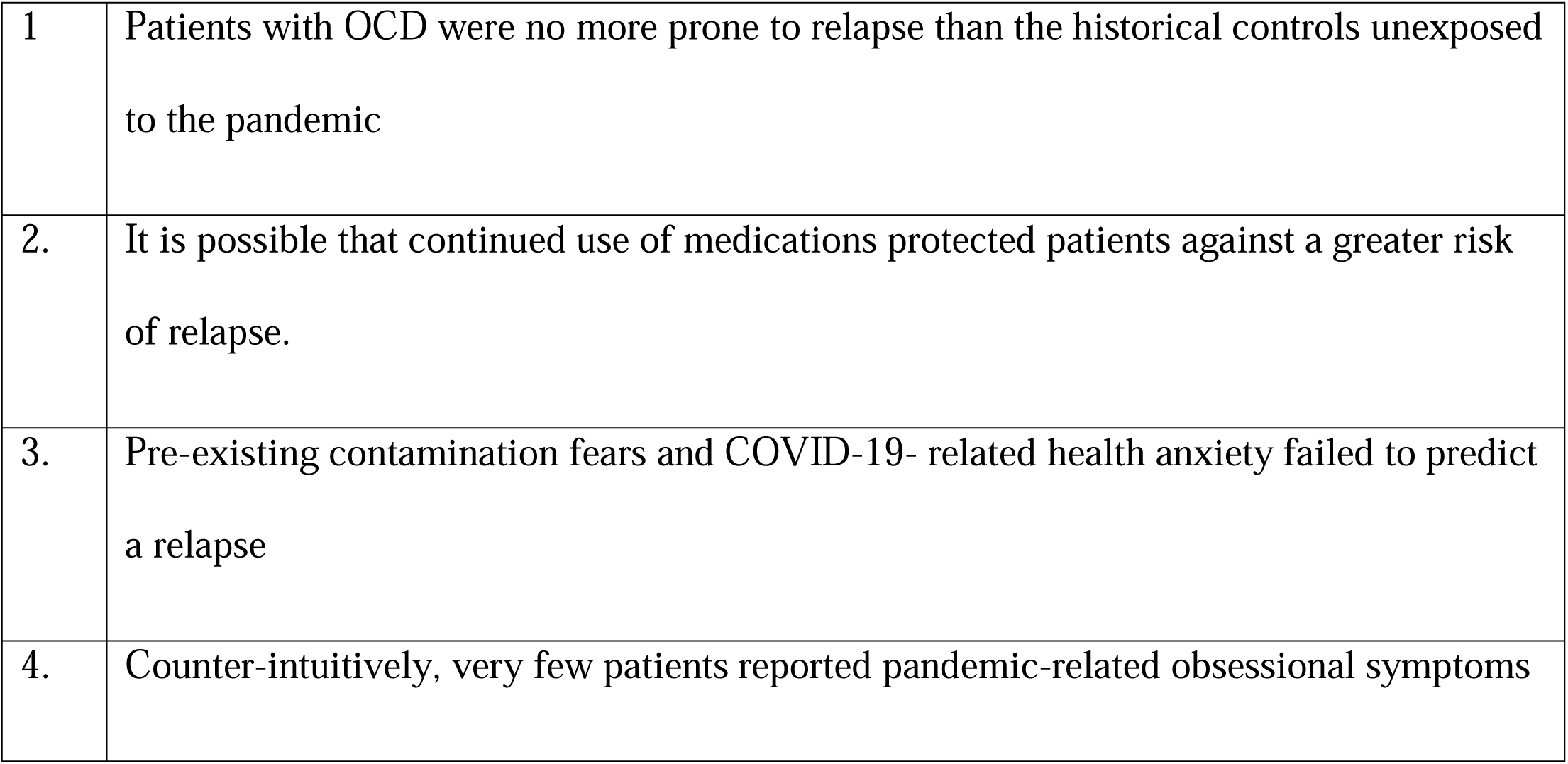

## Introduction

Epidemics can impact people’s mental and emotional health. This has been studied during previous outbreaks of Swine Flu (Wheaton MG, Abramowitz and Berman, NC, Fabricant, LE, Olatunji, 2012), Zika (Blakey and Abramowitz, 2017), and Ebola (Blakey et al., 2015). The ongoing COVID-19 pandemic has demonstrated considerable potential to negatively impact mental health (Galea et al., 2020; Pfefferbaum and North, 2020; Yao et al., 2020). Because of the highly infectious nature of SARS-CoV2 and precautionary measures advocated to limit spread, there has been avid discussion in the news (“Coronavirus: How those obsessed with cleanliness can cope with anxiety, The Indian Express, 2020, “How O.C.D. and Hand-Washing and Coronavirus Collide - The New York Times, 2020, “The hellish side of handwashing: how coronavirus is affecting people with OCD, The Guardian, 2020, “The reality of OCD during the COVID-19 pandemic - The Hindu, 2020) and in scientific journals (Banerjee, 2020; Fineberg et al., 2020; Fontenelle and Miguel, 2020; Santos, 2020; Shafran et al., 2020) on the potential impact of the pandemic on people suffering from obsessive-compulsive disorder (OCD), particularly on how the spike in anxiety about this pandemic could be precipitating and exacerbating OCD (Banerjee, 2020; Fineberg et al., 2020; Fontenelle and Miguel, 2020; French and Lyne, 2020).

This is understandable in the context of safety guidelines that have been widely publicized and discussed (“Advice for public,” WHO, 2020, “How to Protect Yourself & Others | CDC, 2020). These uniformly emphasize the need to avoid potential sources of COVID-19 infection, the importance of social distancing, the use of facial masks, and frequent washing/cleaning in a rather ritualized manner (“Hand Hygiene Recommendations”, CDC, 2020, “Social Distancing, Quarantine, and Isolation,” CDC, 2020). Patients with OCD often employ similar measures to deal with their obsessive fears of contamination and infection. Therefore, the line between adequate safety precautions and compulsive rituals may be blurred for those suffering from OCD. It has been reported, anecdotally, that patients with contamination OCD have expressed doubts about the rationality of ERP during the pandemic (Fineberg et al., 2020).

Anxiety and compensatory behaviors in the context of pandemics have been previously studied in healthy volunteers. One such sample revealed high levels of anxiety during the H1N1 pandemic of 2009–2010; in this study, health anxiety, contamination fears, disgust sensitivity (Wheaton et al., 2012) and pre-existing obsessive-compulsive symptoms (Brand et al., 2013) predicted H1N1-related anxiety. Another study on healthy volunteers during the 2015–2016 Zika virus outbreak found that contamination-related threat estimates significantly predicted Zika virus-related anxiety (Blakey and Abramowitz, 2017). A recent study attempted to identify COVID-19 themed psychopathology from clinical records of 918 patients with a psychiatric diagnosis; however, only 36 of the 918 had a diagnosis of OCD (Rohde et al., 2020).

The only study on the course of OCD during this pandemic is from Italy (Davide et al., 2020). Of the 30 patients studied during the pandemic, 4 prior remitters experienced a relapse of OCD and there was an overall worsening of OCD post-pandemic. Illness exacerbation was associated with an inability to work from home, living with an elderly parent, and the presence of contamination-related symptoms (Davide et al., 2020).

It has been suggested that individuals with OCD may be unable to ‘unlearn’ responses to obsolete threats, and therefore have a greater propensity to prolonged virus-induced distress and anxiety (Fineberg et al., 2020). There have, for example, been reports in the literature, of patients with OCD developing obsessional fears of contamination due to HIV/AIDS (Fisman and Walsh, 1994; Kraus and Nicholson, 1996; Schechter et al., 1991). It is in this context, that it is important to study how the COVID–19 pandemic impacts not only the severity of OCD, but also its symptomatology.

We systematically studied the short-term course of OCD during the COVID-19 pandemic by comparing the relapse rate in the OCD ‘pandemic’ cohort with a group of ‘historical’ OCD controls, evaluated a year before the pandemic. We hypothesized a higher rate of relapse in the OCD ‘pandemic’ cohort compared to the ‘historical’ controls and that relapse in the ‘pandemic’ cohort would be predicted by COVID-19 related health anxiety and pandemic -related dysfunction.

## Method

### Participants

The OCD ‘pandemic’ cohort consisted of a consecutive group of patients with a primary diagnosis of DSM-5 OCD who attended the specialty OCD clinic of the National Institute of Mental Health and Neurosciences (NIMHANS), Bangalore, India for a follow-up visit between 1^st^ October 2019 and 29th February 2020. They were telephonically interviewed over two weeks between 26th April and 12th May 2020, which is approximately 2 months after COVID-19 was declared a pandemic by the World Health Organization(“WHO Director-General’s opening remarks at the media briefing on COVID-19 - 11 May 2020,” WHO, 2020) (see Figure 1 for the recruitment process). This telephonic assessment was done after a mean duration of 3.25 (± 0.81) months following their previous visit to the clinic.

**Figure 1.**
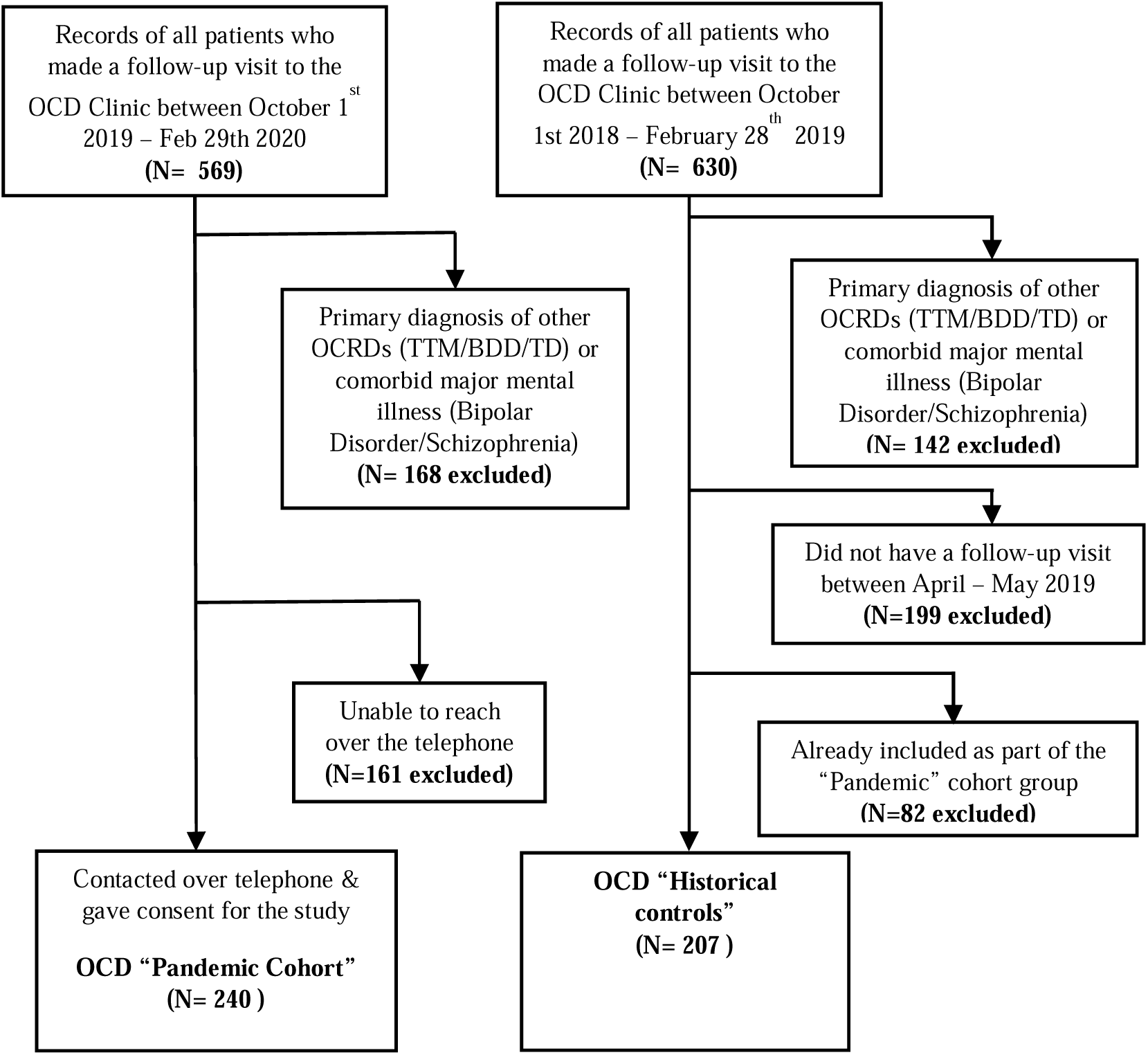
Flow diagram of study recruitment describing the sample ascertainment in the two study groups – the OCD “Pandemic” cohort and the OCD “Historical Control” cohort.

**Figure 2.**
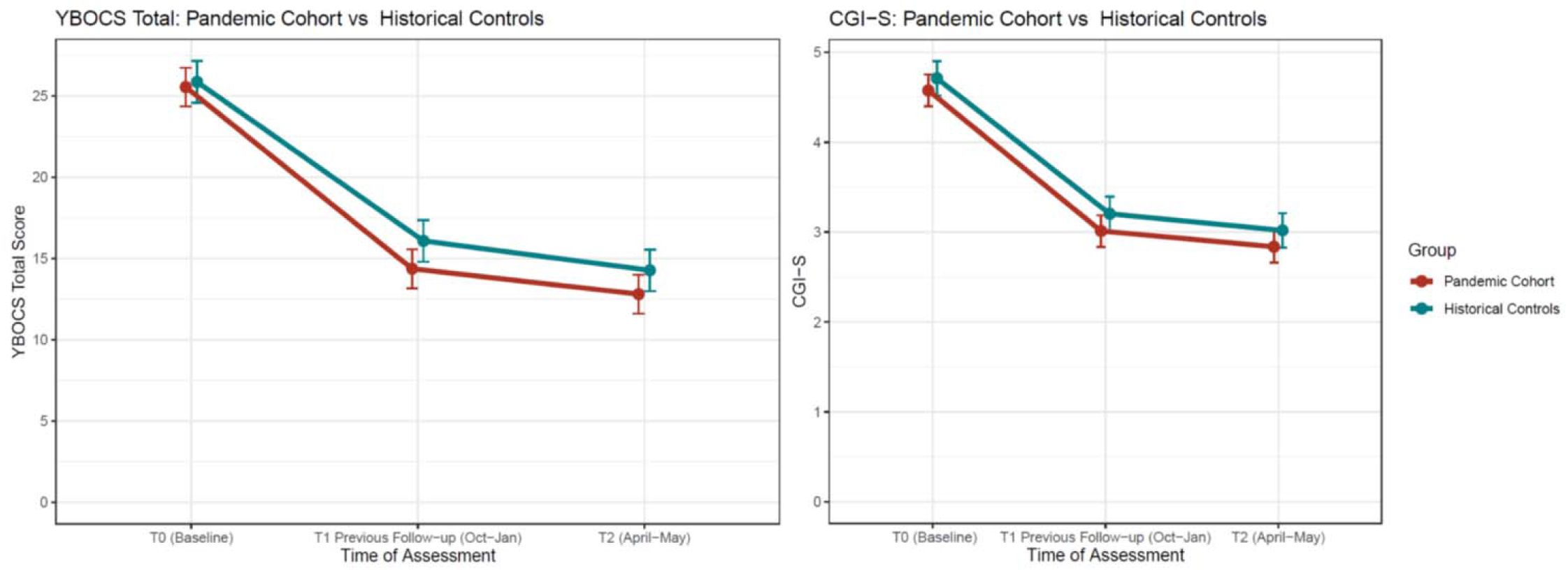
YBOCS Total Score & CGI-S Trajectories in the OCD Pandemic Cohort versus the Historical Control Cohort. YBOCS **–** Yale-Brown Obsessive-Compulsive Scale, CGI-S – Clinical Global Impression (Severity) Scale. T0 (Baseline) the first visit to the clinic, which varies between subjects. T1 indicated the previous follow-up before the pandemic in the pandemic cohort, and the corresponding last follow-up in the historical controls one year prior. T2 indicates the assessment done over a telephonic follow-up in the pandemic cohort, and the corresponding follow-up visit to the clinic for the historical controls

We also reviewed the clinical charts of an independent group of OCD patients (historical controls), who were followed-up at the OCD Clinic, a year before the “pandemic cohort” (i.e., between October 2018 and February 2019), and had another follow-up visit between April and May 2019 in order to compare their relapse rate with that of the pandemic cohort. In all, our sample included 240 pandemic OCD subjects and 207 historical OCD controls (See Figure 1).

### Assessments

For the sake of clarity, the time points of the assessments are designated as follows: “T0” for the baseline assessment (first visit to the OCD clinic, NIMHANS) of both the cohorts, “T1” for the visit between October 2019 and February 2020 for the “pandemic cohort” and October 2018 to February 2019 for the “historical controls”, and “T2” for the telephonic interview for the pandemic cohort and visit between April and May 2019 for the historical controls.

All the OCD patients (pandemic subjects and historical controls) had been previously evaluated at baseline (T0) using the Mini International Neuropsychiatric Interview (MINI; version 6.0.0 till January 2017 and version 7.0.0 from January 2017 onwards) (Sheehan et al., 1998), the Yale-Brown Obsessive-Compulsive Scale (YBOCS) (Goodman et al., 1989) and the Clinical Global Impression - Severity (CGI-S) and improvement (CGI-I) scales (Guy et al, 1976) by trained post- graduate residents in psychiatry and diagnoses were confirmed by a senior clinician of the OCD clinic. The follow-up data of the historical controls (T1 and T2) that included the YBOCS and the CGI ratings was retrieved from the clinical records.

The second follow-up (T2) assessment for the ‘pandemic’ cohort was conducted telephonically. Assessments during the telephonic interview included specific diagnostic evaluations for the major depressive disorder, generalized anxiety disorder and panic disorder using the appropriate sections of the MINI 7.0.0 and also a detailed clinical interview to check if they have developed any new obsessions or compulsions (as defined in the DSM-5) with COVID-19 themes. We also administered the YBOCS (checklist and severity scale), the CGI, the COVID-Threat Scale (CTS) (Wheaton, M.G., Ward, H.E., Sanders, P. R., Reel, J.E., & Van Meter, 2020), and the Work and Social Adjustment Scale (WSAS) (Mundt et al., 2002).

The CTS is used to measure the degree of anxiety related to contracting or spreading COVID-19 (Wheaton, M.G., Ward, H.E., Sanders, P. R., Reel, J.E., & Van Meter, 2020). The scale was translated into five Indian languages (Kannada, Tamil, Telugu, Malayalam, and Hindi), following norms laid out by the World Health Organization (“WHO | Process of translation and adaptation of instruments,” 2010). The questions were read out to patients during the telephonic interview and they were asked to choose their response from the available options.

The WSAS (Mundt et al., 2002) is a measure of functional impairment. Subjects were provided instructions to rate how various aspects of their lives in five domains (work, home management, social leisure, and private leisure, and close relationships) were affected after the pandemic. The original wording of the individual items, which begin with “*Because of my illness*”, was modified to “*Because of the pandemic*”. This scale was also translated with the same procedures as outlined previously. Similar to the CTS, these questions were also read out to patients during the telephonic interview.

The Ethics Committee of the NIMHANS approved the study. Patients of the ‘pandemic’ OCD cohort gave consent orally during the telephonic interview. All the data was anonymized and stored in secure servers.

Outcome criteria were based on the International Consensus Criteria definitions for OCD (Mataix-Cols et al., 2016). Response was defined as having a ≥ 35% reduction in the YBOCS total score along with a CGI-I of 1 or 2. Remission was defined as having a YBOCS total of <12, along with a CGI-S or 1 or 2. We classified patients as responders, non-responders and remitters comparing the YBOCS and CGI scores between “T0” and “T1”. For those patients who were responders at “T1”, we looked for relapse at “T2”. Relapse was defined as “not any longer meeting the criteria for response”, along with a CGI-I of 6 or above. Additionally, we use the term “partial remission” for those who fulfilled criteria for response but not remission at “T1”.

Inter-rater reliability (IRR) exercises for the Y-BOCS ratings are done every three months as part of academic/training activities of the specialty service, using transcripts of YBOCS interviews. The clinicians who evaluated patients in the ‘pandemic’ cohort (LPS, SB, AT, MB, CK and VS) were part of this IRR activity which was done in October 2019, January and April 2020. The intra-class correlation coefficient (ICC) (two-way, fixed raters) for the three IRR exercises ranged between 0.76–0.93 for obsessions, 0.94–0.98 for compulsions, and 0.92–0.97 for the total YBOCS score. Internal consistency (Cronbach’s Alpha) of the telephonic YBOCS ratings was found to be 0.94 for obsessions, and 0.97 for the compulsion sub-scales.

### Statistical Analysis

We calculated the power of our study using the software G*Power version 3.1(Erdfelder et al., 2009). In a previous longitudinal follow-up study in OCD done at our center(Cherian et al., 2014) the rates of relapse within 1-year follow was found to be 13%. Based on z–test of difference in proportions between two independent groups, to detect an increase in relapse rate of 10% (one-tailed) from an expected rate of 13%, with an alpha error probability of 0.05, the power of the study with the current sample size was found to be 0.87.

Change in the YBOCS total score was analyzed using a linear mixed-effect model because of the unbalanced nature of the data (dissimilar sample sizes, non-homoscedasticity i.e., differences in variances both between and within groups at each time point, etc.), which limit the use of the repeated-measures ANOVA. The fixed effects predictors included were: Group (“pandemic” versus “historical” cohort), Time at three levels (T0, T1 and T2), and the Group × Time interaction. We also included a random intercept for time, along with unstructured covariance within subjects, to account for the variability at the baseline and different response patterns at each time point. Any of the baseline characteristics that were found to be significantly different between the two groups were included as covariates in the model. We used the Chi-Square likelihood ratio test to compare between the models for each fixed-effect predictor. The planned post-hoc comparison was the difference in YBOCS total change score between T1 and T2. Tukey’s P-value adjustment method was used and values <0.05 were considered significant.

We used binary logistic regression to compare the relapse rate between the “pandemic” and the “historical” group. The Group (“pandemic” vs “historical”) was included as the main predictor of interest, with other relevant factors, such as partial remission at last follow-up, and medication adherence, as co-variates. It is known that partial remission has been associated with a greater risk of relapse (Cherian et al., 2014; Eisen et al., 2013). Similarly, medication non-adherence may contribute to relapse (Batelaan et al., 2017). A separate binary logistic regression was carried out to identify potential predictors of relapse in the “pandemic” cohort. In this analysis, the CTS total score, the WSAS total score, and other relevant factors such as age, urbanicity (living in a town or city), presence of contamination dimension as a principal symptom, number of failed SRI trials, and medication non-adherence were included as predictors. The CTS and the WSAS were included because we hypothesized them to be predictive of a relapse. We included urbanicity because of a possible higher prevalence of COVID-19 in urban areas(Fortaleza et al., 2020), age because of higher virulence of the virus in the elderly(Zhou et al., 2020), and the contamination dimension because it may confer vulnerability to develop excessive washing or avoidance due to COVID-19(Banerjee, 2020; Fineberg et al., 2020). Number of failed SRI trials indicates degree of resistance to primary pharmacological treatment and may be a potential predictor (Jakubovski et al., 2013). Discontinuation of SRI treatment has also been shown increase rates of relapse (Fineberg et al., 2007).

All analyses were performed with R (version 3.6.2) using the base packages, and the lme4 package for mixed-effects regression.

## Results

### Baseline Characteristics

The subjects included in the pandemic cohort (n=240) did not differ from the subjects who could not be contacted (n=161) with respect to age, sex, age-at-onset, duration of illness, severity of illness as determined by the total YBOCS score and the CGI-S, the response rate at last follow-up before the pandemic (T1) and treatment history (Supplementary Table 1). The pandemic cohort and the historical controls were comparable on all the clinical characteristics (Table 1).

**Table 1.**
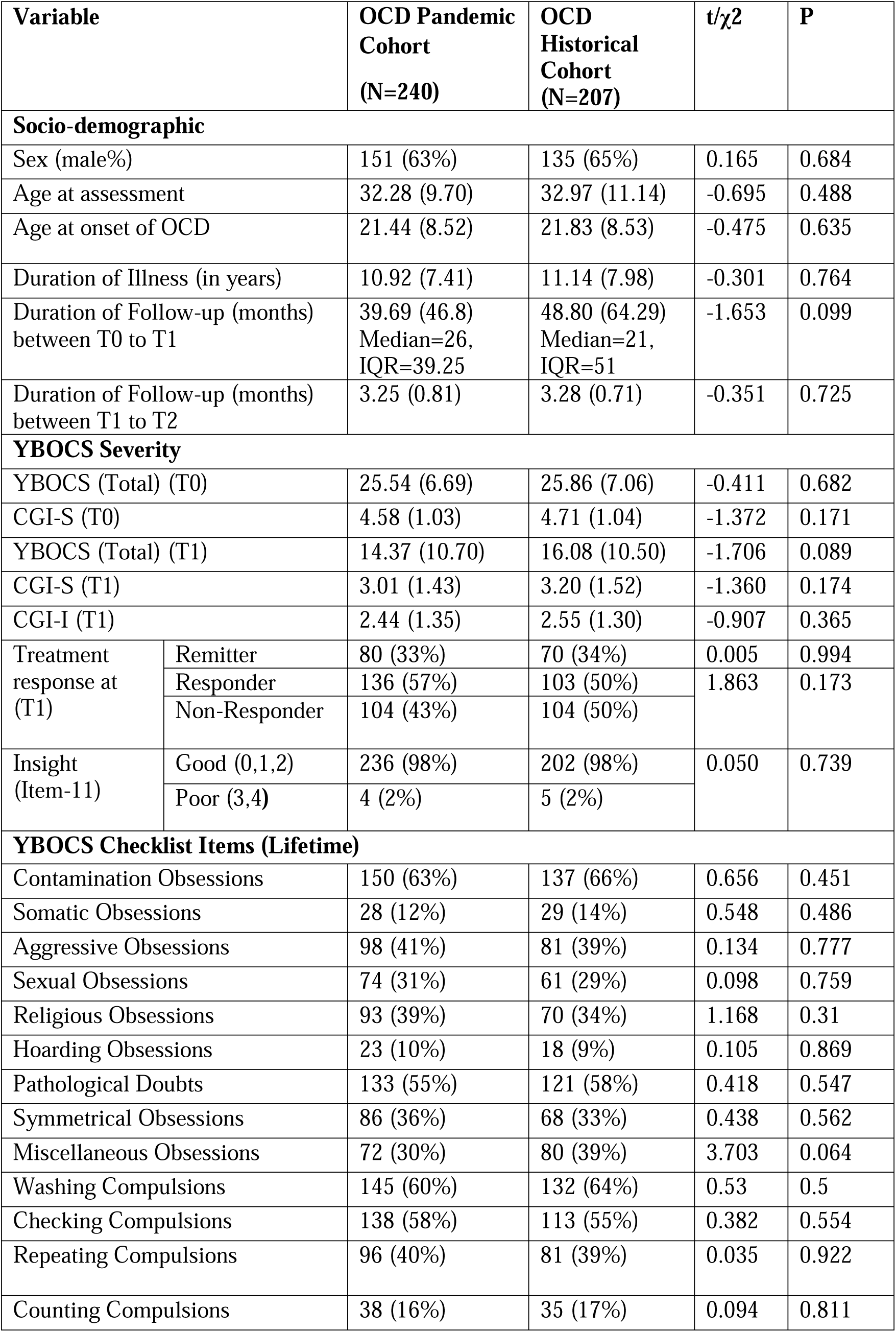
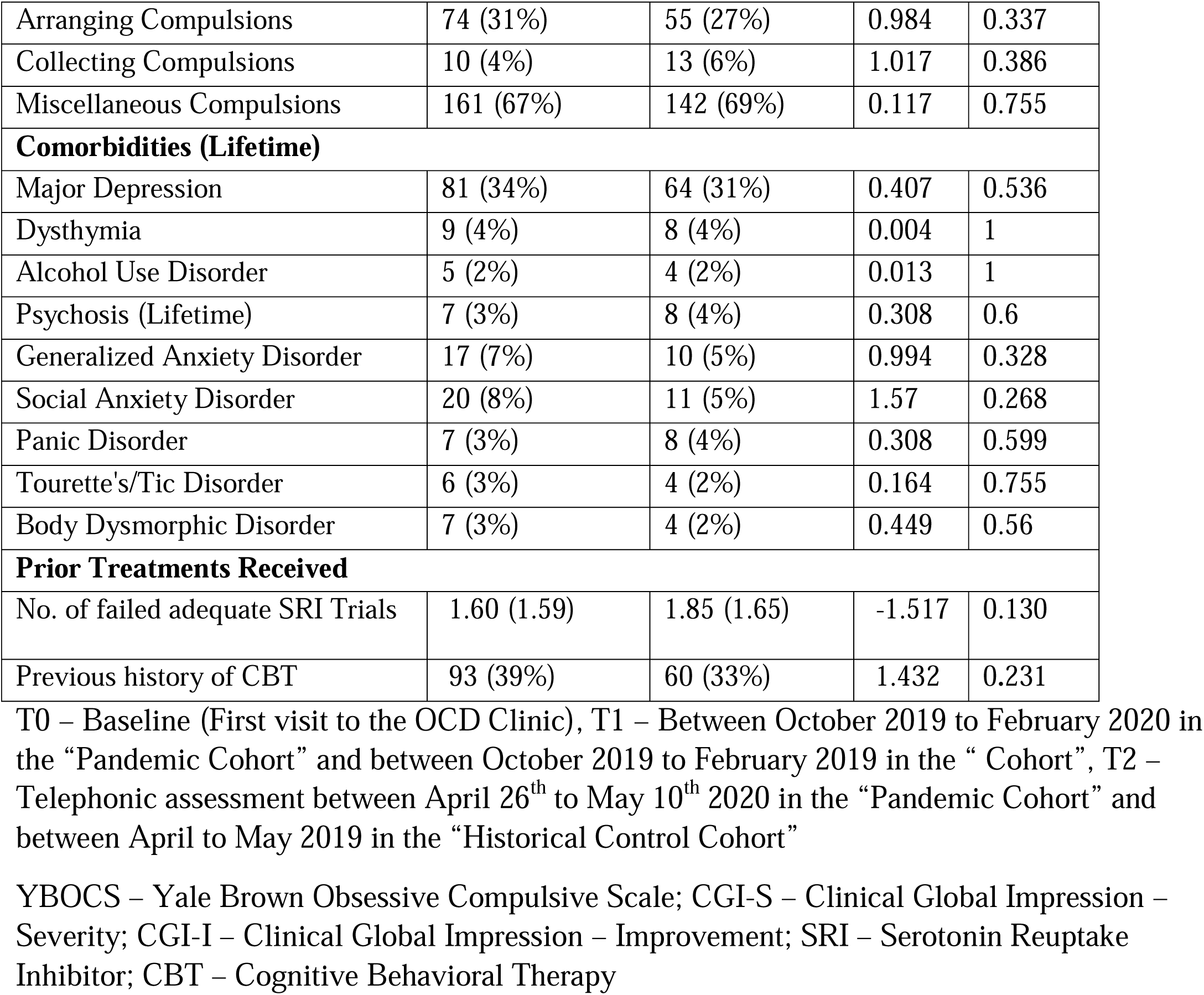
Socio-demographic and Clinical Characteristics of the cohorts.

| Variable | | OCD Pandemic Cohort<br>(N=240) | OCD Historical Cohort<br>(N=207) | t/$\chi^2$ | P |
| --- | --- | --- | --- | --- | --- |
| **Socio-demographic** |  |  |  |  |  |
| Sex (male%) |  | 151 (63%) | 135 (65%) | 0.165 | 0.684 |
| Age at assessment |  | 32.28 (9.70) | 32.97 (11.14) | -0.695 | 0.488 |
| Age at onset of OCD |  | 21.44 (8.52) | 21.83 (8.53) | -0.475 | 0.635 |
| Duration of Illness (in years) |  | 10.92 (7.41) | 11.14 (7.98) | -0.301 | 0.764 |
| Duration of Follow-up (months) between T0 to T1 |  | 39.69 (46.8)<br>Median=26,<br>IQR=39.25 | 48.80 (64.29)<br>Median=21,<br>IQR=51 | -1.653 | 0.099 |
| Duration of Follow-up (months) between T1 to T2 |  | 3.25 (0.81) | 3.28 (0.71) | -0.351 | 0.725 |
| **YBOCS Severity** |  |  |  |  |  |
| YBOCS (Total) (T0) |  | 25.54 (6.69) | 25.86 (7.06) | -0.411 | 0.682 |
| CGI-S (T0) |  | 4.58 (1.03) | 4.71 (1.04) | -1.372 | 0.171 |
| YBOCS (Total) (T1) |  | 14.37 (10.70) | 16.08 (10.50) | -1.706 | 0.089 |
| CGI-S (T1) |  | 3.01 (1.43) | 3.20 (1.52) | -1.360 | 0.174 |
| CGI-I (T1) |  | 2.44 (1.35) | 2.55 (1.30) | -0.907 | 0.365 |
| Treatment response at (T1) | Remitter | 80 (33%) | 70 (34%) | 0.005 | 0.994 |
|  | Responder | 136 (57%) | 103 (50%) | 1.863 | 0.173 |
|  | Non-Responder | 104 (43%) | 104 (50%) |  |  |
| Insight (Item-11) | Good (0,1,2) | 236 (98%) | 202 (98%) | 0.050 | 0.739 |
|  | Poor (3,4) | 4 (2%) | 5 (2%) |  |  |
| **YBOCS Checklist Items (Lifetime)** |  |  |  |  |  |
| Contamination Obsessions |  | 150 (63%) | 137 (66%) | 0.656 | 0.451 |
| Somatic Obsessions |  | 28 (12%) | 29 (14%) | 0.548 | 0.486 |
| Aggressive Obsessions |  | 98 (41%) | 81 (39%) | 0.134 | 0.777 |
| Sexual Obsessions |  | 74 (31%) | 61 (29%) | 0.098 | 0.759 |
| Religious Obsessions |  | 93 (39%) | 70 (34%) | 1.168 | 0.31 |
| Hoarding Obsessions |  | 23 (10%) | 18 (9%) | 0.105 | 0.869 |
| Pathological Doubts |  | 133 (55%) | 121 (58%) | 0.418 | 0.547 |
| Symmetrical Obsessions |  | 86 (36%) | 68 (33%) | 0.438 | 0.562 |
| Miscellaneous Obsessions |  | 72 (30%) | 80 (39%) | 3.703 | 0.064 |
| Washing Compulsions |  | 145 (60%) | 132 (64%) | 0.53 | 0.5 |
| Checking Compulsions |  | 138 (58%) | 113 (55%) | 0.382 | 0.554 |
| Repeating Compulsions |  | 96 (40%) | 81 (39%) | 0.035 | 0.922 |
| Counting Compulsions |  | 38 (16%) | 35 (17%) | 0.094 | 0.811 |

|  |  |  |  |  |
| --- | --- | --- | --- | --- |
| Arranging Compulsions | 74 (31%) | 55 (27%) | 0.984 | 0.337 |
| Collecting Compulsions | 10 (4%) | 13 (6%) | 1.017 | 0.386 |
| Miscellaneous Compulsions | 161 (67%) | 142 (69%) | 0.117 | 0.755 |
| **Comorbidities (Lifetime)** |  |  |  |  |
| Major Depression | 81 (34%) | 64 (31%) | 0.407 | 0.536 |
| Dysthymia | 9 (4%) | 8 (4%) | 0.004 | 1 |
| Alcohol Use Disorder | 5 (2%) | 4 (2%) | 0.013 | 1 |
| Psychosis (Lifetime) | 7 (3%) | 8 (4%) | 0.308 | 0.6 |
| Generalized Anxiety Disorder | 17 (7%) | 10 (5%) | 0.994 | 0.328 |
| Social Anxiety Disorder | 20 (8%) | 11 (5%) | 1.57 | 0.268 |
| Panic Disorder | 7 (3%) | 8 (4%) | 0.308 | 0.599 |
| Tourette's/Tic Disorder | 6 (3%) | 4 (2%) | 0.164 | 0.755 |
| Body Dysmorphic Disorder | 7 (3%) | 4 (2%) | 0.449 | 0.56 |
| **Prior Treatments Received** |  |  |  |  |
| No. of failed adequate SRI Trials | 1.60 (1.59) | 1.85 (1.65) | -1.517 | 0.130 |
| Previous history of CBT | 93 (39%) | 60 (33%) | 1.432 | 0.231 |

T0 – Baseline (First visit to the OCD Clinic), T1 – Between October 2019 to February 2020 in the “Pandemic Cohort” and between October 2019 to February 2019 in the “ Cohort”, T2 – Telephonic assessment between April 26<sup>th</sup> to May 10<sup>th</sup> 2020 in the “Pandemic Cohort” and between April to May 2019 in the “Historical Control Cohort”
YBOCS – Yale Brown Obsessive Compulsive Scale; CGI-S – Clinical Global Impression – Severity; CGI-I – Clinical Global Impression – Improvement; SRI – Serotonin Reuptake Inhibitor; CBT – Cognitive Behavioral Therapy

Nearly all patients were on active pharmacological treatment in both the OCD groups, details of which are provided in Table 2. Additionally, most patients in each sample were on stable medication, and more than 80% of patients in both groups reported full adherence to medication.

**Table 2.** Ongoing pharmacological treatment in the samples.

| Treatment | | OCD<br>Pandemic<br>Cohort<br>(N=240) | OCD Historical<br>Cohort<br>(N=207) | F/$\chi^2$ | P |
| --- | --- | --- | --- | --- | --- |
| Current SRI<br>Trial<br>[mean dose ±<br>SD, mg] | Fluoxetine [67.5 ± 13.3] | 61 (25.4%) | 62 (29.9%) | 0.929 | 0.335 |
| | Escitalopram [23.8 ± 6.8] | 61 (25.4%) | 43 (20.7%) | 1.315 | 0.252 |
| | Sertraline [201 ± 55.3] | 52 (21.7%) | 30 (14.5%) | 3.355 | 0.067 |
| | Fluvoxamine [253 ± 80] | 19 (7.9%) | 27 (13.1%) | 2.633 | 0.105 |
| | Paroxetine [60.2 ± 19.5] | 10 (4.2%) | 6 (2.9%) | 0.216 | 0.642 |
| | Clomipramine [184 ±<br>48.9] | 12 (5%) | 24 (11.6%) | 5.667 | 0.017 |
| | Venlafaxine [228 ± 31] | 18 (7.5%) | 12 (5.8%) | 0.278 | 0.598 |
|  | Not on SRI | 7 (2.9%) | 3 (1.5%) | 0.526 | 0.468 |
| Mean duration of current SRI trial (weeks) |  | 72.49<br>(87.88),<br>Median=36,<br>IQR=20-96 | 81.57 (118.4)<br>Median=32<br>IQR=16-80 | 1.558 | 0.122 |
| Stable SRI Medication (12 weeks) |  | 222 (95.3%) | 187 (91.7%) | 1.802 | 0.179 |
| Current<br>Augmentation<br>Trial<br>[mean dose ±<br>SD, mg] | Risperidone [2.4 ± 1.2] | 22 (9.2%) | 29 (14.0%) | 2.122 | 0.145 |
| | Aripiprazole [11.3 ± 6.5] | 28 (11.7%) | 33 (15.9%) | 1.380 | 0.240 |
| | Haloperidol [1.5 ± 1.4] | 2 (0.8%) | 0 | 0.367 | 0.545 |
| | Olanzapine [6.25 ± 2.5] | 2 (0.8%) | 0 | 0.367 | 0.545 |
| | Quetiapine [171 ± 95.4] | 3 (1.3%) | 3 (1.4%) | <0.001 | 1.000 |
| | Memantine [23.3 ± 8.2] | 4 (1.7%) | 2 (1%) | 0.053 | 0.818 |
| | N-Acetyl Cysteine<br>[1800±0] | 0 (0%) | 1 (0.5%) | 0.005 | 0.941 |
| | Clomipramine [75±0] | 2 (0.8%) | 0 | 0.367 | 0.545 |
| | Amisulpride [250±57.7] | 3 (1.3%) | 1 (0.5%) | 0.126 | 0.723 |
|  | No augmentation | 173 (72.1%) | 134 (66.7%) | 2.459 | 0.117 |
| Mean duration of current augmentation trial (weeks) |  | 63.25<br>(85.44),<br>Median=36,<br>IQR=19.5-54.5 | 43.26 (61.25)<br>Median=20,<br>IQR=12-49.5 | 1.558 | 0.122 |
| Stable augmentation (12 weeks) |  | 62 (92.5%) | 61 (83.6%) | 1.864 | 0.172 |
| Medication<br>adherence<br>during the<br>interval period<br>(T1 to T2) | Adherent | 224 (93.3%) | 195 (94.2%) | 0.033 | 0.855 |
|  | Discontinued for >2<br>weeks | 16 (6.7%) | 12 (5.8%) |  |  |
| CBT practice (among those<br>who earlier received it) | Yes | 53 (57%) | 26 (43.3%) | 2.723 | 0.138 |
|  | No | 40 (43%) | 34 (56.7%) |  |  |

SRI – Serotonin Reuptake Inhibitor; CBT – Cognitive Behavioral Therapy

### Comparison of Illness severity between the pandemic and historical cohorts

Comparisons between the two OCD groups concerning outcomes are shown in Table 3. The linear mixed-effect model showed a significant main effect of time. There was an overall mean reduction in YBOCS total score, of 9.88 (95% CI 8.59 – 11.17) at previous follow up (T1), and 11.70 (95% CI 10.41 - 12.99) at the “Current” time point (T2) (Type II Wald Chi-Square = 922.83, p<0.001). The effect of the Group*Time interaction was not found to be significant (Type II Wald Chi-Square Likelihood ratio= 2.73, p=0.255), and the mean change in YBOCS score from the T1 to T2 was around the same in both the groups (Table 3). Figure 1 shows the plotted trajectories comparing the two groups.

**Table 3.**
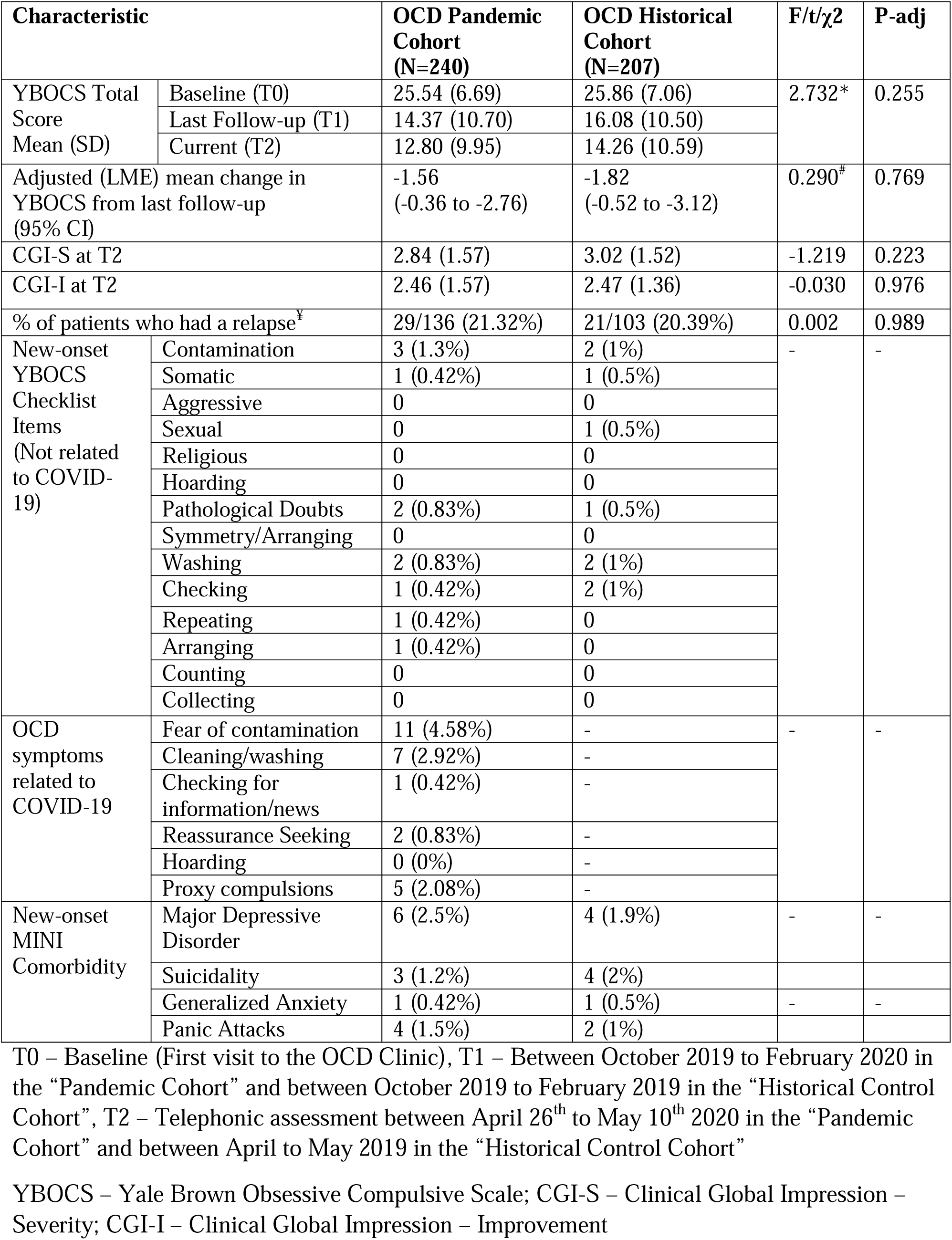
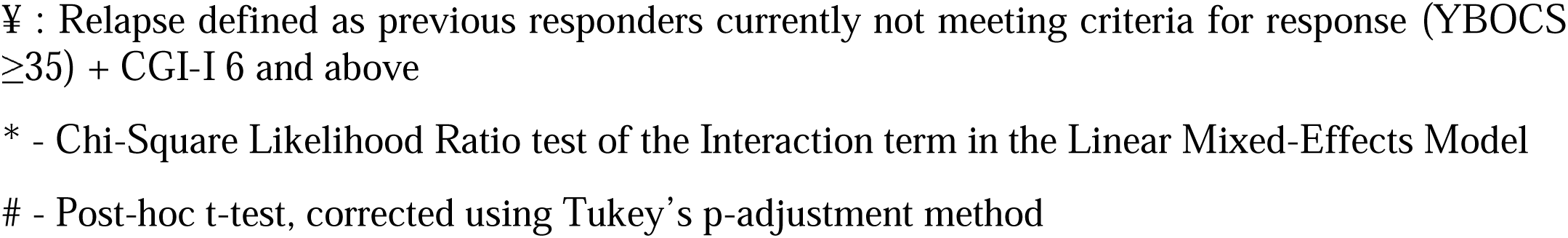
Comparisons of Outcomes between the two OCD Cohorts.

### Comparison of relapse rate between the pandemic and historical cohorts

Around 20% of subjects relapsed in both the cohorts (Table-3). Binary logistic regression was done to compare relapse rates between groups, adjusting for partial remission and for medication non-adherence (which was dichotomously coded as “Full”, or “Non-adherent”). Adjusted odds of relapse due to the pandemic was found to be 0.81 (95% CI 0.41 -1.59, p=0.535). Significantly high odds of ratios were found for the co-variates “partial remission” – 4.01 (95% CI 2.04 – 8.19, p<0.001), and “non-adherence” 6.83 (95% CI 2.39 - 20.29, p<0.001).

### New-Onset OC symptoms

Table 3 also shows the rates of new-onset OC symptoms and comorbidity in the two OCD cohorts. There were very few patients who endorsed new symptoms in the YBOCS checklist, in either group. Also, only a small proportion (overall less than 6%) in the “pandemic” cohort reported obsessions or compulsions related to COVID-19. The number of patients who reported new-onset comorbidity between T1 and T2 assessments was also small.

### Predictors of Relapse

Table 4 shows the results of the logistic regression analysis done to find the association between relevant clinical and contextual factors and relapse amongst patients in the “pandemic” cohort. As seen, relapse following the pandemic was significantly predicted only by “Partial Remission” status at last follow-up, and by WSAS total scores. The other key variables, including principal symptom contamination/washing or the CTS total score (for COVID-19 related anxiety), were not found to be significant predictors.

**Table 4.** Predictors of “Relapse” (n=29) in the “Pandemic Cohort” among the Responders (n=136)

|  | **B (SE)** | **z value** | **Odds Ratio (95% CI of OR)** | **P** |
| --- | --- | --- | --- | --- |
| Intercept | -1.86 (1.33) | -1.39 | 0.16 (0.01 - 1.97) | 0.164 |
| Age in years | -0.04 (0.03) | -1.27 | 0.96 (0.9 - 1.02) | 0.205 |
| Urbanicity | -0.77 (0.57) | -1.34 | 0.46 (0.15 - 1.43) | 0.179 |
| Partial Remission in last follow-up (0=No, 1=Yes) | 1.55 (0.58) | 2.69 | 4.72 (1.6 - 15.82) | 0.007 |
| Principal Symptom Contamination/Washing | 0.25 (0.55) | 0.45 | 1.28 (0.42 - 3.81) | 0.652 |
| Number of Failed SRI Trials | 0.3 (0.17) | 1.75 | 1.35 (0.97 - 1.91) | 0.080 |
| Medication Non-Adherence | 1.01 (0.76) | 1.33 | 2.74 (0.58 - 12.05) | 0.185 |
| COVID-related anxiety (CTS – Total) | -0.06 (0.04) | -1.42 | 0.94 (0.87 - 1.02) | 0.157 |
| Work & Social Mal-Adjustment following the Pandemic (WSAS) | 0.17 (0.04) | 4.17 | 1.19 (1.1 - 1.3) | <0.001 |

Prediction Accuracy of the model = 88.3%; Chi-square=61.814; p.value=<0.001;
Nagelkerke $R^2$ (Cragg and Uhler) = 0.589
SRI – Serotonin Reuptake Inhibitor, CTS – COVID Threat Scale, WSAS – Work & Social Adjustment Scale

## Discussion

We systematically evaluated the impact of the COVID–19 pandemic on OCD in a large sample of patients with OCD who were mostly stabilized on treatment, with over half having responded to medication. The main findings of our study are i) OCD subjects did not appear to have any worsening in the severity of illness during the pandemic compared to historical controls during the same period in the previous year; ii) Relapse rates among the ‘pandemic’ OCD group are similar to those in the ‘historical’ control group; and iii) Very few patients developed COVID–19 related obsessive-compulsive symptoms.

Our study findings are counterintuitive and reassuring, since most OCD subjects on stable medication seem to have withstood the impact of COVID–19, at least in the short term. It was thought that individuals with OCD would be more significantly and directly affected by the COVID-19 outbreak than those with other psychiatric disorders (Fineberg et al., 2020). The spike in anxiety due to the virus and the elaborate safety guidelines issued by many national and international agencies emphasizing on frequent washing and cleaning (in a rather ritualized manner) was thought to be the perfect recipe for worsening of pre–existent contamination fears or for the onset of new contamination fears related to contracting COVID–19. Experts from the International College of Obsessive-Compulsive Spectrum Disorders (ICOCS) and the Obsessive-Compulsive and Related Disorders Research Network of the European College of Neuropsychopharmacology (OCRN) have recently issued guidelines on how to manage OCD during the times of COVID–19 (Fineberg et al., 2020). However, in contrast to speculation in the scientific community and the lay media that suggest a higher risk of relapse in those suffering from OCD, it is interesting to note that our cohort of OCD patients did not have a significantly higher relapse rate during the pandemic than the group of historical controls. Incomplete remission before the pandemic and pandemic–related dysfunction as indicated by higher scores on the WSAS, and not the fear of COVID–19 per se (measured by the CTS) predicted relapse. Incomplete/partial remission has been associated with relapse in previous studies of naturalistic outcome of OCD (Cherian et al., 2014).

There are several potential reasons for a comparable rate of relapse in the pandemic group with that in the historical controls. Firstly, medication may have had a protective effect against a relapse. Secondly, the relapse rate during the pandemic is not higher than in controls possibly because of a relatively shorter duration of exposure to the pandemic. Thirdly, it is also possible that the OCD subjects were not exposed to COVID-19 stimuli because of extensive and strict lockdown and safety precautions; these may have perhaps neutralized their fears, resulting in a lesser propensity for relapse or worsening.

Our findings have important clinical implications. Most of our patients were on stable doses of SRIs and/or augmenting agents and were largely adherent to treatment. It implies that continued treatment with medications may have prevented worsening/relapse during the pandemic and lockdown. In the background of continued and understandable concern about the possibility of OCD worsening due to the pandemic, our study offers some solace in the fact that SRIs may have a protective role against a relapse/worsening of the severity of illness. We of course do not have a control group who were not on medications, but clinicians may consider advising people who suffer from OCD not to discontinue or reduce the dose of SRIs. This is particularly important now because getting help from CBT therapists may not be all that easy due to varied degrees of lockdown and continued emphasis on social distancing. Although our study does suggest that continued treatment with medications may have played a role in preventing the worsening of illness severity, we do not know if continued treatment with CBT alone will have a similar effect considering the challenges involved in recommending exposure and response prevention and the blurred line between ‘rational’ concerns and ‘OCD concerns’.

### Strengths and Limitations

There are several strengths to this study. Our sample was largely on stable medications in the preceding 12 weeks, which suggests that treatment changes did not confound our results. Sample size was reasonably large and all the participants were assessed using standard tools. The availability of a historical control group followed up during the same period, the previous year (i.e., 2018–2019) helped ascertain that relapses in the OCD pandemic group were perhaps not due to COVID–19 but a function of the natural course of the illness.

Findings of our study have to be interpreted in the light of some obvious limitations. Although the patients whom we could not assess were no different from the ones we could, it is quite possible that a bias may have crept in to the response pattern. Because of the strict lockdown, we performed assessments telephonically; it is possible that some of the responses on measures such as the CTS and the WSAS may have been influenced by the pattern of reading out the items and recording the responses. The YBOCS assessments may not have been affected much by the method of interviewing since the instrument is clinician–administered and the patients had been previously exposed to the measure and were familiar with it. Although the treatment adherence rate is high, we could not corroborate this by interviewing a relative or caregiver, which is the usual practice in our clinic. We assessed patients about 2 months after declaration of the pandemic by the WHO which is perhaps a relatively short period to study its impact.

In summary, the course of OCD, at least in the short term, does not appear to be significantly different from the usual course of illness in our sample. It is heartening that relapse rates have not increased in the context of this pandemic. These findings must be interpreted in light of the limitations stated above. Following up on our OCD ‘pandemic’ cohort over this year may help us understand the long-term impact of COVID–19.

## Data Availability

Requests to share anonymized datasheets may be addressed to the corresponding author (SB)

## Acknowledgments

We offer our special thanks to Dr. Abhishek Allam, Prof. PSVN Sharma, Prof. Hemavathi Balachander, Dr. Lekhansh Shukla, Dr. Madhuri HN, and Dr. Karthik S for helping with translation & back translation of scales. We are immensely grateful to the staff at the Medical Records Department, NIMHANS for their efforts in aiding us in Data Collection. We also thank Dr. Neha Kulkarni and Dr. Aishwarya Sachdeva for helping us with data entry. We thank Prof. Jagadisha Thirthalli for his valuable inputs regarding improving the study design and statistical analysis.

---

**Supplementary Table 1.**
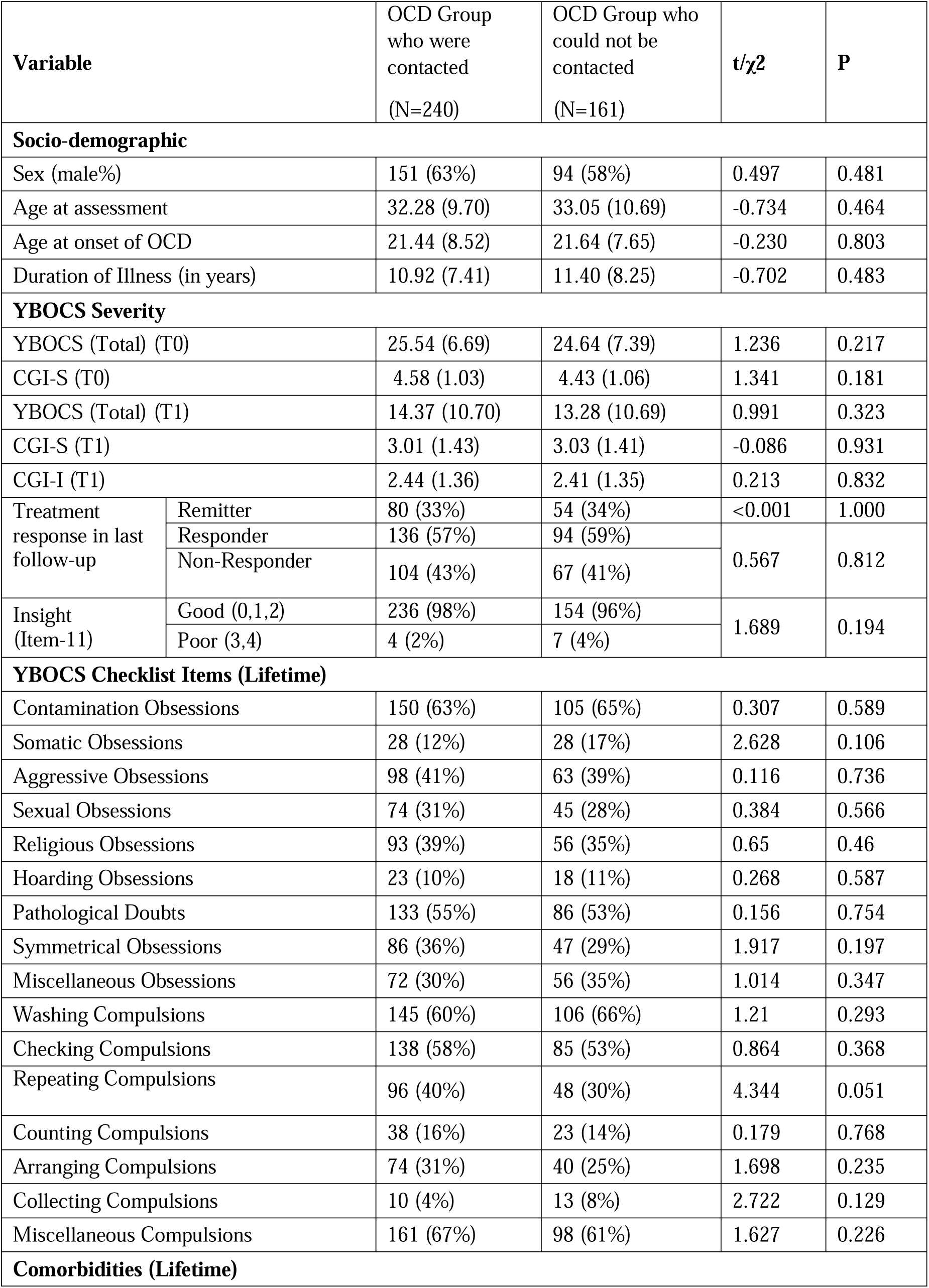
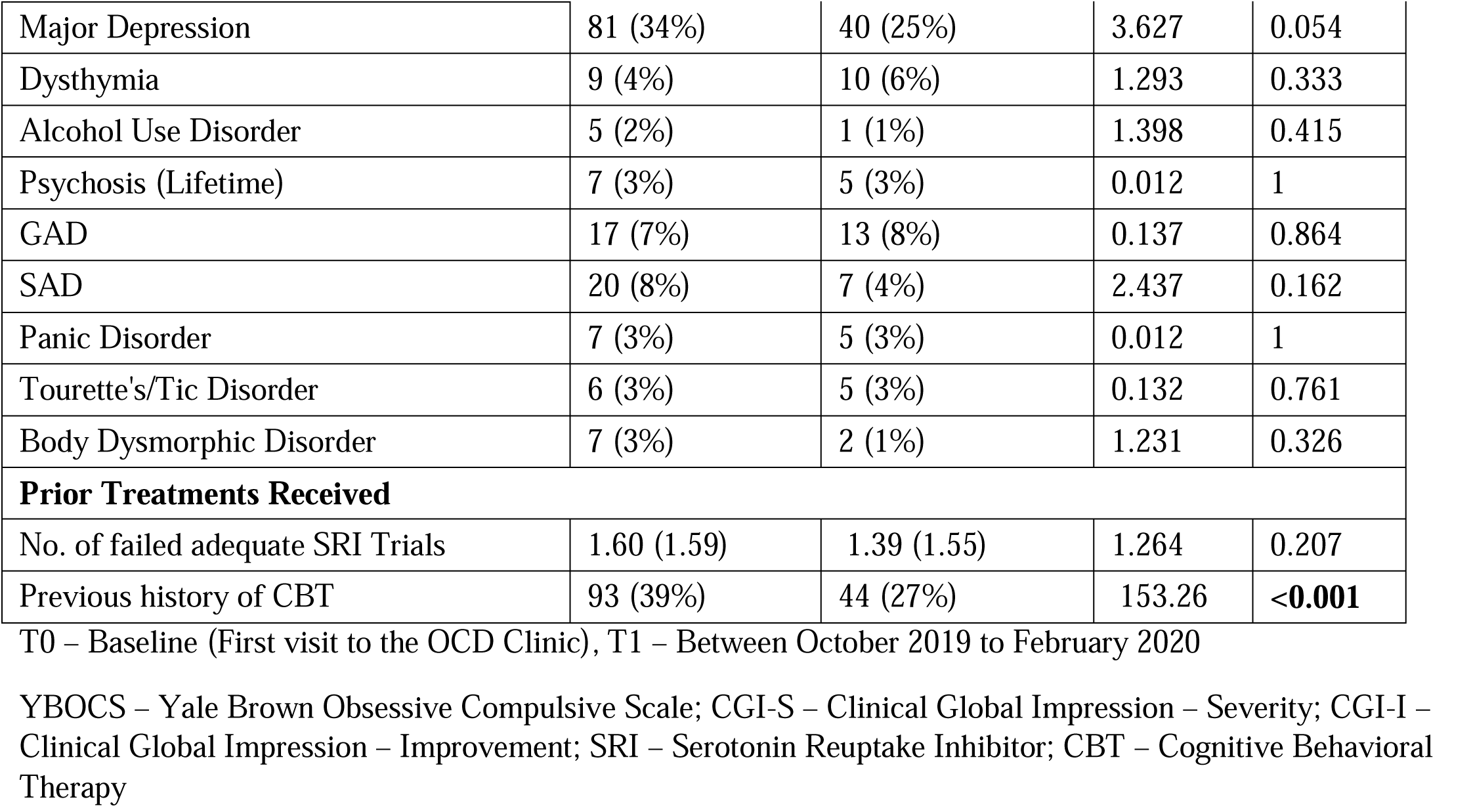
Comparison of Socio-demographic and Clinical Characteristics between those who were contracted versus those who could not be contacted

| Variable | | OCD Group<br>who were<br>contacted<br>(N=240) | OCD Group who<br>could not be<br>contacted<br>(N=161) | t/$\chi^2$ | P |
| --- | --- | --- | --- | --- | --- |
| Socio-demographic |  |  |  |  |  |
| Sex (male%) |  | 151 (63%) | 94 (58%) | 0.497 | 0.481 |
| Age at assessment |  | 32.28 (9.70) | 33.05 (10.69) | -0.734 | 0.464 |
| Age at onset of OCD |  | 21.44 (8.52) | 21.64 (7.65) | -0.230 | 0.803 |
| Duration of Illness (in years) |  | 10.92 (7.41) | 11.40 (8.25) | -0.702 | 0.483 |
| YBOCS Severity |  |  |  |  |  |
| YBOCS (Total) (T0) |  | 25.54 (6.69) | 24.64 (7.39) | 1.236 | 0.217 |
| CGI-S (T0) |  | 4.58 (1.03) | 4.43 (1.06) | 1.341 | 0.181 |
| YBOCS (Total) (T1) |  | 14.37 (10.70) | 13.28 (10.69) | 0.991 | 0.323 |
| CGI-S (T1) |  | 3.01 (1.43) | 3.03 (1.41) | -0.086 | 0.931 |
| CGI-I (T1) |  | 2.44 (1.36) | 2.41 (1.35) | 0.213 | 0.832 |
| Treatment<br>response in last<br>follow-up | Remitter | 80 (33%) | 54 (34%) | <0.001 | 1.000 |
|  | Responder | 136 (57%) | 94 (59%) | 0.567 | 0.812 |
|  | Non-Responder | 104 (43%) | 67 (41%) |  |  |
| Insight<br>(Item-11) | Good (0,1,2) | 236 (98%) | 154 (96%) | 1.689 | 0.194 |
|  | Poor (3,4) | 4 (2%) | 7 (4%) |  |  |
| YBOCS Checklist Items (Lifetime) |  |  |  |  |  |
| Contamination Obsessions |  | 150 (63%) | 105 (65%) | 0.307 | 0.589 |
| Somatic Obsessions |  | 28 (12%) | 28 (17%) | 2.628 | 0.106 |
| Aggressive Obsessions |  | 98 (41%) | 63 (39%) | 0.116 | 0.736 |
| Sexual Obsessions |  | 74 (31%) | 45 (28%) | 0.384 | 0.566 |
| Religious Obsessions |  | 93 (39%) | 56 (35%) | 0.65 | 0.46 |
| Hoarding Obsessions |  | 23 (10%) | 18 (11%) | 0.268 | 0.587 |
| Pathological Doubts |  | 133 (55%) | 86 (53%) | 0.156 | 0.754 |
| Symmetrical Obsessions |  | 86 (36%) | 47 (29%) | 1.917 | 0.197 |
| Miscellaneous Obsessions |  | 72 (30%) | 56 (35%) | 1.014 | 0.347 |
| Washing Compulsions |  | 145 (60%) | 106 (66%) | 1.21 | 0.293 |
| Checking Compulsions |  | 138 (58%) | 85 (53%) | 0.864 | 0.368 |
| Repeating Compulsions |  | 96 (40%) | 48 (30%) | 4.344 | 0.051 |
| Counting Compulsions |  | 38 (16%) | 23 (14%) | 0.179 | 0.768 |
| Arranging Compulsions |  | 74 (31%) | 40 (25%) | 1.698 | 0.235 |
| Collecting Compulsions |  | 10 (4%) | 13 (8%) | 2.722 | 0.129 |
| Miscellaneous Compulsions |  | 161 (67%) | 98 (61%) | 1.627 | 0.226 |
| Comorbidities (Lifetime) |  |  |  |  |  |

|  |  |  |  |  |
| --- | --- | --- | --- | --- |
| Major Depression | 81 (34%) | 40 (25%) | 3.627 | 0.054 |
| Dysthymia | 9 (4%) | 10 (6%) | 1.293 | 0.333 |
| Alcohol Use Disorder | 5 (2%) | 1 (1%) | 1.398 | 0.415 |
| Psychosis (Lifetime) | 7 (3%) | 5 (3%) | 0.012 | 1 |
| GAD | 17 (7%) | 13 (8%) | 0.137 | 0.864 |
| SAD | 20 (8%) | 7 (4%) | 2.437 | 0.162 |
| Panic Disorder | 7 (3%) | 5 (3%) | 0.012 | 1 |
| Tourette's/Tic Disorder | 6 (3%) | 5 (3%) | 0.132 | 0.761 |
| Body Dysmorphic Disorder | 7 (3%) | 2 (1%) | 1.231 | 0.326 |
| **Prior Treatments Received** |  |  |  |  |
| No. of failed adequate SRI Trials | 1.60 (1.59) | 1.39 (1.55) | 1.264 | 0.207 |
| Previous history of CBT | 93 (39%) | 44 (27%) | 153.26 | **<0.001** |

T0 – Baseline (First visit to the OCD Clinic), T1 – Between October 2019 to February 2020
YBOCS – Yale Brown Obsessive Compulsive Scale; CGI-S – Clinical Global Impression – Severity; CGI-I – Clinical Global Impression – Improvement; SRI – Serotonin Reuptake Inhibitor; CBT – Cognitive Behavioral Therapy

